# Prospective Cohort Study Identifies Barriers to Point-of-care Ultrasound use in an Academic Emergency Department

**DOI:** 10.1101/2025.08.21.25334181

**Authors:** Rebecca G Theophanous, Alyssa Platt, Timothy Peterson, Maragatha Kuchibhatla, Erica Peethumnongsin

**Affiliations:** Department of Emergency Medicine; Duke University School of Medicine, Durham, NC 27710, USA; Department of Biostatistics and Bioinformatics, Duke University School of Medicine, Durham, NC 27710, USA

## Abstract

**Background/objectives:** Point-of-care ultrasound (POCUS) improves patient care by expedited diagnosis and safer procedures. Despite POCUS benefits, some clinicians, including emergency physicians, do not readily use POCUS. The study objective identified barriers to clinical POCUS use and performed an intervention to address barriers and increase clinical POCUS use.

**Methods:** A prospective cohort study at a single academic hospital included ED attendings, residents, and advance practice providers (APPs). Participants were surveyed on perceived POCUS use barriers (primary outcome) and clinical POCUS were numbers recorded from July 2023-June 2024. A multi-faceted intervention from December 2023-January 2024 addressed identified barriers and involved: in-person POCUS education during shift by ultrasound faculty, clinical POCUS workflow demonstration during resident conference/faculty meetings, and QR code reference files on machines. Secondary outcomes were POCUS workflow knowledge exam scores, clinical ED POCUS scans performed, and revenue. Pre-/post-intervention analysis was performed using independent t-tests.

**Results:** 42/104 ED providers (40.4%) responded to surveys pre-intervention and 28 post-intervention (28.3%). 56 physicians/APPs participated in the in-person POCUS intervention (17 attendings, 34 residents, 5 APPs). Perceived POCUS barriers were time constraints on shift; internet/connectivity problems and losing saved images; forgetting to finish exam worksheets online; images not uploading into Butterfly cloud by the end of shift; and residents performing “phantom scans”. Mean knowledge scores were 10/11 (89%), with 26/28 participants passing (score >90%). Participant self-perceived comfort in performing diagnostic and procedural POCUS increased (p=0.100, p=0.784 respectively). Procedural teaching comfort increased (p=0.022) but not diagnostic teaching (p=0.166). POCUS scan numbers increased from 1511 to 2130 (p=0.0035). Monthly POCUS revenue increased ~$55K in total billed (p=0.0439) and $10K reimbursed (p=0.1225).

**Conclusion:** Identified barriers were incorporated into a multi-faceted approach to improve clinical POCUS workflow processes, with increased ED POCUS use and revenue post-intervention. Future individualized interventions for low POCUS users and institutional initiatives with clinical champions can be studied for improved patient care.

## Background

Point-of-care ultrasound (POCUS) is a valuable diagnostic bedside tool for clinical patient care.^1,2^ Emergency ultrasound advances clinical diagnostics at the bedside, allows for repeat assessments during resuscitation of acutely ill patients, and guides high risk procedures and treatment for improved patient safety.^3,4^ POCUS expedites patient care workflows by reducing emergency department (ED) radiology study orders and transport times, thereby reducing patient disposition times.^1,5^ This yields cost efficiencies and improved ED and hospital throughput. Performing POCUS also improves patient-centered outcomes and satisfaction, through increased time at the bedside and direct patient interaction.^1,5^

The American College of Emergency Physicians (ACEP) and American College of Graduate Medical Education (ACGME) guidelines define the knowledge and performance of twelve core POCUS diagnostic exams and ultrasound-guided procedures as a critical aspect of emergency physician training.^3,4^ Despite increasing POCUS utilization at medical centers nationwide, there persists a significant gap between POCUS training and actual clinical use, including in emergency medicine (EM).^6–9^ Prior studies in the Veterans Affairs (VA) healthcare system and rural EDs have cited multiple factors contributing to this gap in POCUS use, including lack of POCUS training, lack of a systemwide POCUS documentation and archiving program, lack of hospital resources or funding, and for some sites a lack of equipment.^7,8,10–15^ Additional studies have investigated whether creation of an institution-specific POCUS program can increase POCUS utilization in EDs or inpatient services, however this model may not be generalizable to other sites, and sustainability is difficult to achieve.^16–20^

## Objective

Our aim for this study was to identify barriers and facilitators to POCUS use by ED physicians and APPs, including faculty attendings and residents, in a single large academic hospital. We sought to better understand the problems with our ED POCUS clinical system to help overcome these barriers for POCUS use, documentation, archiving, and billing processes. By performing this study, our goal was to increase clinical POCUS use in the department and disseminate the data collected as a model for other EDs or hospital systems.

## Methods

### Study design

We performed a prospective cohort study from July 2023 to June 2024 to identify barriers and facilitators to clinical POCUS use by EM attending physicians, advanced practice providers (APPs), or EM resident physicians currently working at a single academic hospital ED. The study was performed according to the Strengthening the Reporting of Observational Studies in Epidemiology (STROBE) guidelines.^21^ The study was approved by the Institutional Review Board (Pro00114618) and followed ethics guidelines.

### Study setting

The study was conducted at a single tertiary care, academic, level 1 trauma center with 1,048 inpatient hospital beds, almost 85,000 annual ED visits, and 29.8% admissions. The ED has seven diagnostic ultrasound machines, which include two General Electric Healthcare Venue cart-based systems, three Sonosite M-Turbos, and two Sonosite Edges (with two new Sonosite X-porte machines added in July 2024). The ED also has three vascular Sonosite Nanomaxx machines for procedural use and four Butterfly IQ handheld ultrasounds that are currently only used for educational purposes, (although they have the capability and access to be used for clinical scans). The study site has one Emergency Ultrasound Director and Ultrasound Fellowship Director. The EM program has 12 residents per academic year and one emergency ultrasound fellow. Epic Maestrocare is the electronic health record, and Butterfly Network cloud is the ultrasound middleware software for POCUS image storage and archival. Quality assurance (QA) is performed by ultrasound-fellowship trained faculty on a weekly basis for all clinical POCUS scans.

### Selection of Participants

All providers working in the ED at the time of the study were invited to participate. The study sample included 49 attending physicians, 19 APPs, and 36 residents (PGY1-3). We used convenience sampling and recruited participants via email, at EM resident conference, and at monthly EM faculty meetings. ED nurses, technicians, and residents working in the ED from other medical services were excluded as this study focused on physicians (and APPs) trained in ED POCUS use. Per survey demographics, there were 22 male participants, 19 female, 6 Hispanic/Latin/x/Spanish origin, 32 non-Hispanic/Latin/x/Spanish, with mean age 36 [27-56 years], and a mean 8.5 years [0.5-32 years] of clinical work in EM. Eleven participants reported fellowship training. Follow up was not required after the study.

### Data collection and outcomes

All ED faculty, APPs, and residents were emailed a secured RedCap survey (with a website link and QR code) pre- and post-intervention to assess their understanding of the ED clinical POCUS workflow and asked their opinions on barriers or potential facilitators for the process. (Supplement 1) The survey was created by ultrasound-content experts and based on validated surveys from other POCUS studies.^16,22^

Based on the pre-intervention survey results, we modified an educational POCUS workflow intervention on best practice, systematic clinical ultrasound performance, documentation, and archival workflow practice. ED ultrasound faculty then reeducated participants on the clinical POCUS workflow via an intervention. First, we created an online presentation and reference document outlining the standardized POCUS image acquisition and archival process. This was emailed to providers, uploaded to the secured ED clinical folder, and accessible via new QR codes attached to the ED ultrasound machines. Second, we performed an in-person demonstration of the clinical ED POCUS process during shift changes in the ED over a 4-week period for EM attendings, APPs, and residents. This was conducted live by ED ultrasound faculty. Finally, we reviewed monthly ED clinical POCUS billing data and collected data on the number and type of POCUS scans done in the department to assess the effectiveness of our intervention. We surveyed ED attendings and residents via RedCap 6-months post-intervention including a knowledge exam to obtain feedback on the POCUS intervention and assess efficacy and workflow improvements. The goal of these process interventions was to improve clinical workflow processes and not to cause harm to participants or their employment.

### Study outcomes

The primary outcome was the identified barriers and facilitators to POCUS use at our site. Additional measures included participant self-perceived POCUS use, comfort in performing and teaching ultrasound scans, and a knowledge assessment on clinical POCUS workflow from pre- and post-intervention survey data.

Secondary outcomes for assessing the impact of the POCUS program intervention were the number and type of clinical POCUS exams performed by ED physicians, APPs, and residents in the 6-month periods before and after the intervention. We hypothesized that clinical ED POCUS use would increase post-intervention. We also collected data on ED POCUS billing and reimbursement.

### Data sources

We used Epic MaestroCare for patient chart review and Butterfly Network cloud software to review POCUS scans done in the emergency department. We used our institution’s secured RedCap server for participant surveys and obtained ED clinical POCUS billing data from administrative and support staff for review.

### Bias

The survey questionnaire was written by the study PI and edited by the ED ultrasound faculty group prior to distribution so that it would be institution-specific and to reduce bias. The survey was pilot tested with an ED physician at another local institution with research training who was not eligible for the study.

Participants were enrolled with implied consent after reading the study information sheet with the RedCap survey link emailed by the study PI. All participation was voluntary, and participants could withdraw at any time from the study. All survey data was secured in an institutional database per standard research protocols, and only study personnel had access to data. Participant names and dates were necessary to link pre/post-intervention survey data to assess for improvement after the POCUS intervention, and for analysis of responses to understand barriers, for scientific integrity, and to maximize process improvements. Survey data was deidentified by the study PI and coded with a key for biostatistician review and analysis. POCUS studies are saved within the institution’s approved and secured Butterfly cloud system and written reports in Epic Maestrocare as done in standard clinical care. Only deidentified data was presented to the ED group for feedback and process improvements.

We used Kirkpatrick’s levels of learning to evaluate our POCUS intervention, assessing participants’ reaction to the POCUS intervention through survey questionnaires and their learning through a knowledge assessment on clinical POCUS workflow. We evaluated behavior through self-perceived comfort in performing and teaching POCUS and via in-person observed demonstrations of faculty completing a clinical POCUS scan during on-shift teaching. Finally, program results were measured through cost-benefit analysis and overall conclusions made about the program.^23^

### Statistical methods

Pre-/post-intervention analysis from survey data was performed using independent t-tests. POCUS barriers and facilitators were descriptively reported. For the impact evaluation, secondary outcomes included the number and type of clinical ED POCUS scans performed. Monthly and aggregate pre-/post-intervention proportions of ED patients undergoing each type of ultrasound scan were calculated and compared using independent t-tests. Data is also presented on ED POCUS billing and reimbursement. P-values were calculated using an alpha of 0.05 for statistical significance. Missing data from blank responses on questionnaires was reported in the tables. Participants lost to follow-up (e.g. did not participate in the intervention or post-questionnaire) could not be incorporated into the analysis or results. Statistical analysis was performed with the SAS 9.4 statistical software (SAS Institute Inc., Cary, NC).

Sample size: As this was a pilot descriptive study identifying barriers and facilitators to POCUS use, sample size and power are not reported.

## Results

### Characteristics of the study subjects

In the 6-month pre-intervention to 6-month post-intervention period from July 1, 2023 to June 30, 2024, a total of 81,649 patients were seen in the ED and 3641 POCUS were performed. Of 104 total ED providers, 42 responded to the questionnaire prior to the intervention (40.4%), including 26 EM attendings, 14 EM residents, 1 APP, and 1 unidentified user. The 5 EM attendings acting as study researchers abstained from voting to prevent bias and are not included. 28 participants responded to the questionnaire post intervention (28.3%). 56 participated in the in-person POCUS intervention (17 faculty, 34 residents, 5 APPs). Participant characteristics and questionnaires are included in Table 1. The study timeline is presented in Figure 1.

**Table 1:** Participant Characteristics of ED providers from pre/post-course questionnaire responses.

|  | **Pre-intervention survey participants, n=42** | **Post-intervention survey participants, n=28** |
| --- | --- | --- |
| **Age (mean (S.D.) median (IQR) [range in years])** | 36 (7.96)<br>34 (8)<br>[27-56]<br>[Missing 3] | 36 (6.82)<br>34.5 (8.5)<br>[27-56]<br>[Missing 0] |
| **Sex** | 22 male, 19 female<br>[Missing 2] | 15 male, 13 female<br>[Missing 0] |
| **Ethnicity** | N=6<br>Hispanic/Latin/x/Spanish origin, n=32 non-Hispanic/Latin/x/Spanish origin<br>[Missing 5] | N=5<br>Hispanic/Latin/x/Spanish origin, n=21 non-Hispanic/Latin/x/Spanish origin, n=2 other<br>[Missing 0] |
| **Race** | N=9 Asian, n=4 Black or African American, n=24 white, n=0 Native Hawaiian or Other Pacific Islander, n=0 American Indian or Alaska Native, n=2 other<br>[Missing 2] | N=6 Asian, n=0 Black or African American, n=21 white, n=0 Native Hawaiian or Other Pacific Islander, n=0 American Indian or Alaska Native, n=1 other<br>[Missing 0] |
| **Training level** | N=27 attending physicians, n=1 APP, n=14 EM residents (n=3 PGY1, n=5 PGY2, n=6 PGY3)<br>[Missing 0] | N=17 attending physicians, n=2 APPs, n=9 EM residents (n=3 PGY1, n=5 PGY2, n=1 PGY3)<br>[Missing 0] |
| **Fellowship training (yes)\*** | N=11 | N=9 |
| **Years working in the ED (mean (S.D.) median (IQR) [range in years])** | 8.6 (8.65)<br>8.4 (7.5)<br>[0.5-32]<br>[Missing 1] | 8.3 (6.53)<br>8 (9)<br>[2-35]<br>[Missing 0] |
| **Age range per category** | n=7, age 30-40<br>n=8, age 41-50<br>n=9, age 51-60+<br>[Missing 3] | n=8, age 30-40<br>n=7, age 41-50<br>n=6, age 51-60+<br>[Missing 0] |
| **Number of POCUS personally performed in residency training** | n=5, 0-50 POCUS<br>n=3, 51-100 POCUS<br>n=8, 101-150 POCUS<br>n=27, >150 POCUS<br>[Missing 0] | n=0, 0-50 POCUS<br>n=4, 51-100 POCUS<br>n=3, 101-150 POCUS<br>n=21, >150 POCUS<br>[Missing 0] |
| **Current clinical ED POCUS use** | n=23, use POCUS often (>1 time per week)<br>n=16, use POCUS somewhat often (1-2 times per month)<br>n=2, occasional POCUS use | n=24, use POCUS often (>1 time per week)<br>n=4, use POCUS somewhat often (1-2 times per month)<br>n=0, occasional POCUS |
| | ($<1$ time per month)<br>n=1, rarely (1-2 times per year)<br>n=0 never use POCUS<br>[Missing 1] | use ($<1$ time per month)<br>n=0, rarely (1-2 times per year)<br>n=0 never use POCUS<br>[Missing 0] |
| **Number and training level of POCUS in-person teaching intervention participants** | N=56 participants (n=17 faculty, n=11 PGY3, n=11 PGY2, n=12 PGY1, n=5 APPs) |  |

\*Fellowships completed: health services research, emergency medical services, sports medicine, hyperbaric and undersea medicine, geriatric emergency medicine, emergency ultrasound

**Figure 1.**
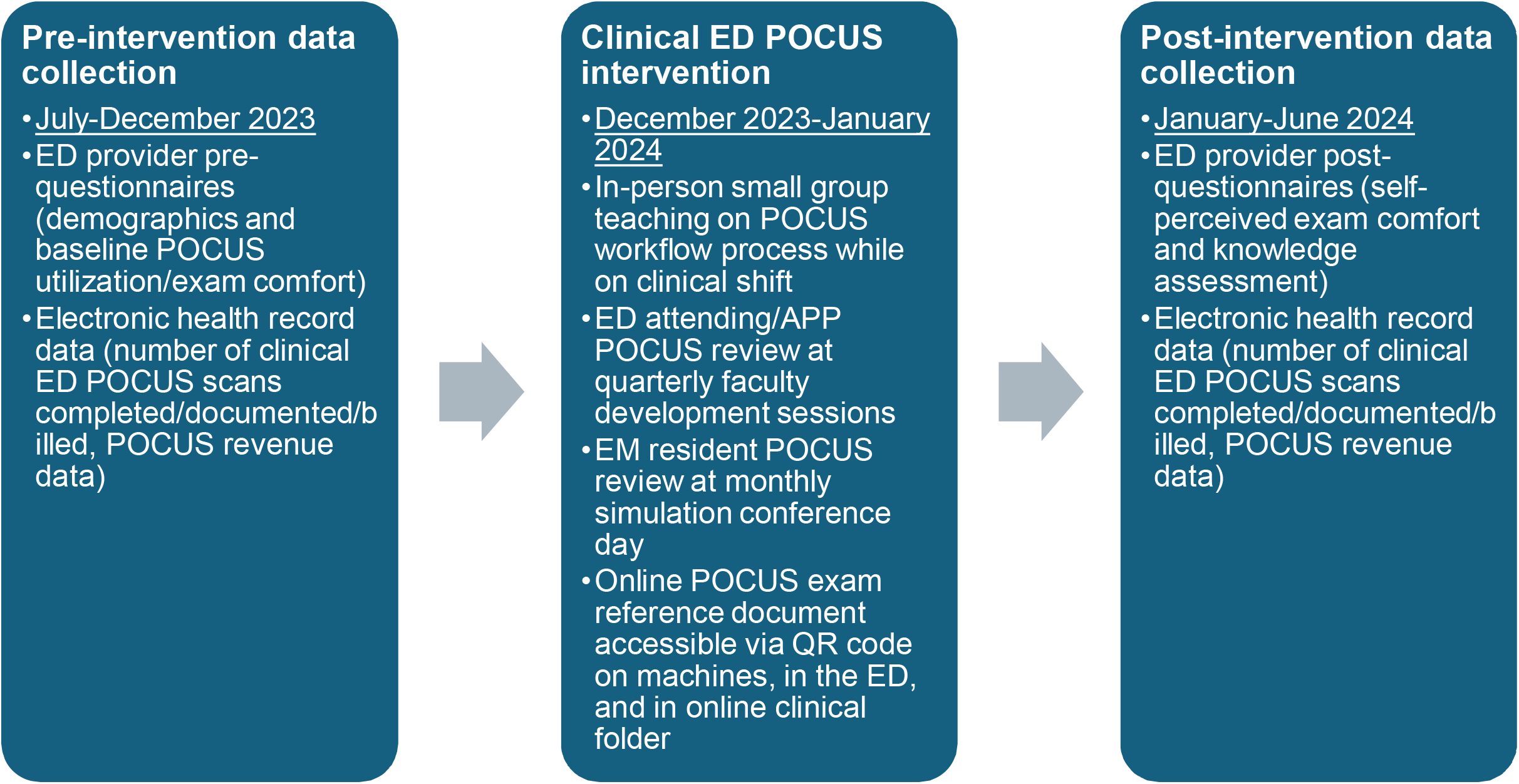
Clinical ED POCUS workflow process improvement timeline

Per survey demographics, there were 22 male participants, 19 female, 6 Hispanic/Latin/x/Spanish origin, 32 non-Hispanic/Latin/x/Spanish, with mean age 36 [27-56 years], and a mean 8.5 years of clinical work in EM [0.5-32 years]. Eleven participants reported fellowship training. Responses per level of training included: 1 APP (2.4%), 3 PGY1 residents (7.1%), 5 PGY2 (11.9%), 6 PGY3 (14.3%), no Fellows, and 27 attending physicians (64.3%).(Table 1)

The most common perceived POCUS barriers were *time constraints on shift* (30 or 69.8% respondents reported moderate/significant barrier); *internet or connectivity problems and losing saved images* (26 or 61.9% respondents); *forgetting to complete the Butterfly cloud worksheet after scan completion, images not uploaded in Butterfly cloud by the end of the shift* (both had 24 or 55.8% respondents reporting moderate/significant barrier); *difficulty in locating the ultrasound machines* (20 or 46.6% respondents); *not knowing how to use the machine tracker system* (18 or 41.8% respondents); *residents are performing “phantom scans” and not saving their images* (19 or 46.3% respondents); *and problems with the machine malfunctioning* (21 or 48.9% respondents). Some respondents felt that *residents* or *APPs not completing worksheets in Butterfly* was a barrier (15 or 36.6% respondents) and that *faculty were not signing the completed studies* (14 or 34.2% respondents). The least common perceived barriers were: *not knowing how to document an ultrasound scan; not knowing how to use the Butterfly cloud archiving software; losing access to a non-associated scan after the two-week time window when patients are removed from the list; lack of confidence in performing or obtaining POCUS images; ready availability of radiology ultrasound; and hospital credentialing not including all necessary specific POCUS exams*.(Table 2a, Figure 2a)

**Table 2a:** POCUS Barriers and facilitators identified from participant surveys.

| **Clinical POCUS Barriers:** | **Clinical POCUS Facilitators:** |
| --- | --- |
| Time constraints on shift (30/42)<br>Internet or connectivity problems and losing saved images (26/42)<br>Forgetting to fill out the worksheet on Butterfly cloud after completing the scan (24/42)<br>Images or study not being uploaded into Butterfly cloud by the end of the shift (24/42)<br>Not being able to locate the ultrasound machines (20/42)<br>Not knowing how to use the machine tracker system (18/42)<br>Residents are performing “phantom scans” and not saving their images (19/42)<br>Problems with the ultrasound machine malfunctioning (21/42)<br>Residents or APPs not completing worksheets in Butterfly (15/42)<br>Faculty were not signing the completed studies (14/42) | Clear documentation protocols (30/42)<br>Hands-on teaching sessions (24/42)<br>*Incentives for completing scans (24/42)<br>Additional online training (13/42)<br>Penalties for not completing a certain number of POCUS scans (9/42) |

\*Incentives: competition, monetary bonus for faculty, more stringent faculty credentialing process)

**Figure 2.**
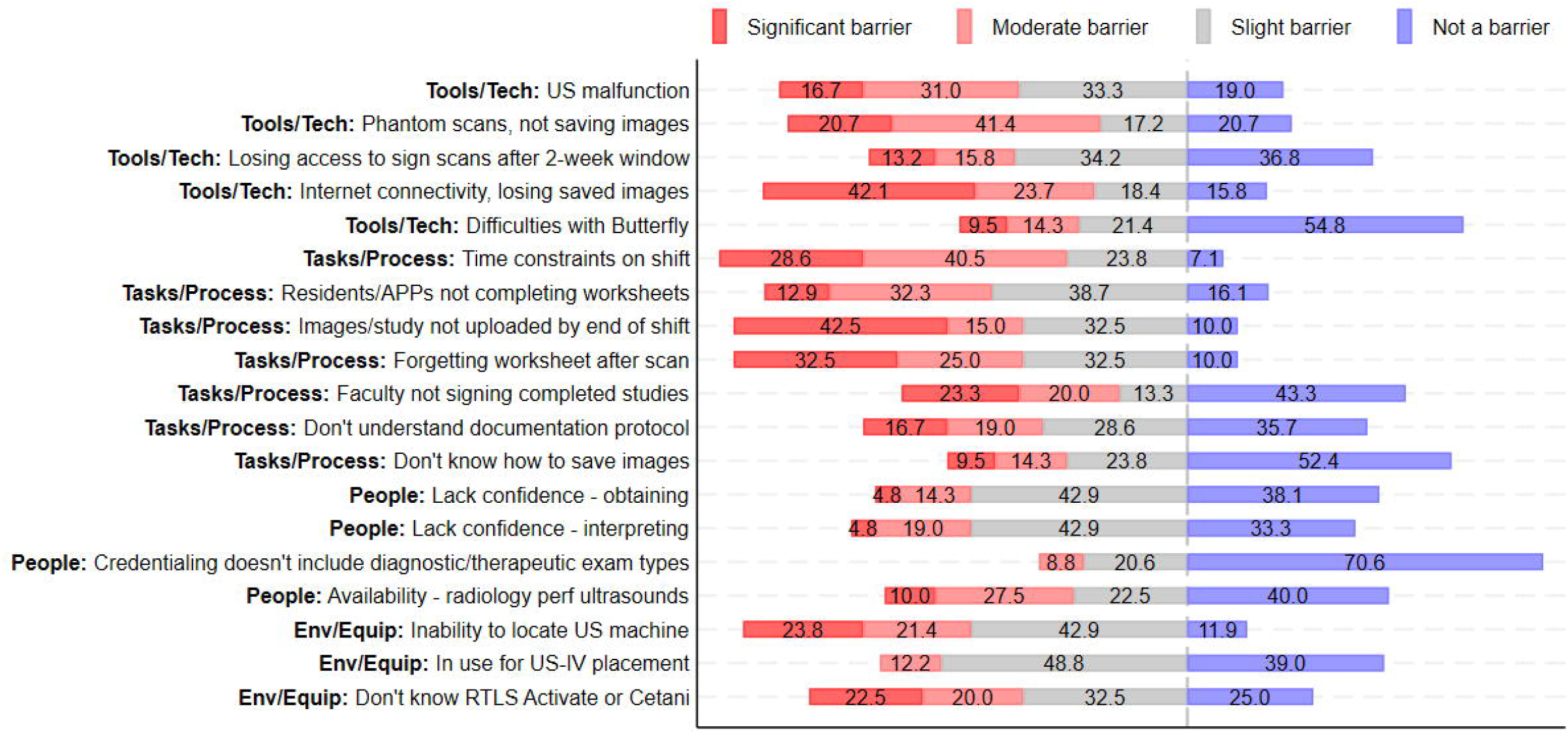
POCUS Barriers

**Figure 3.**
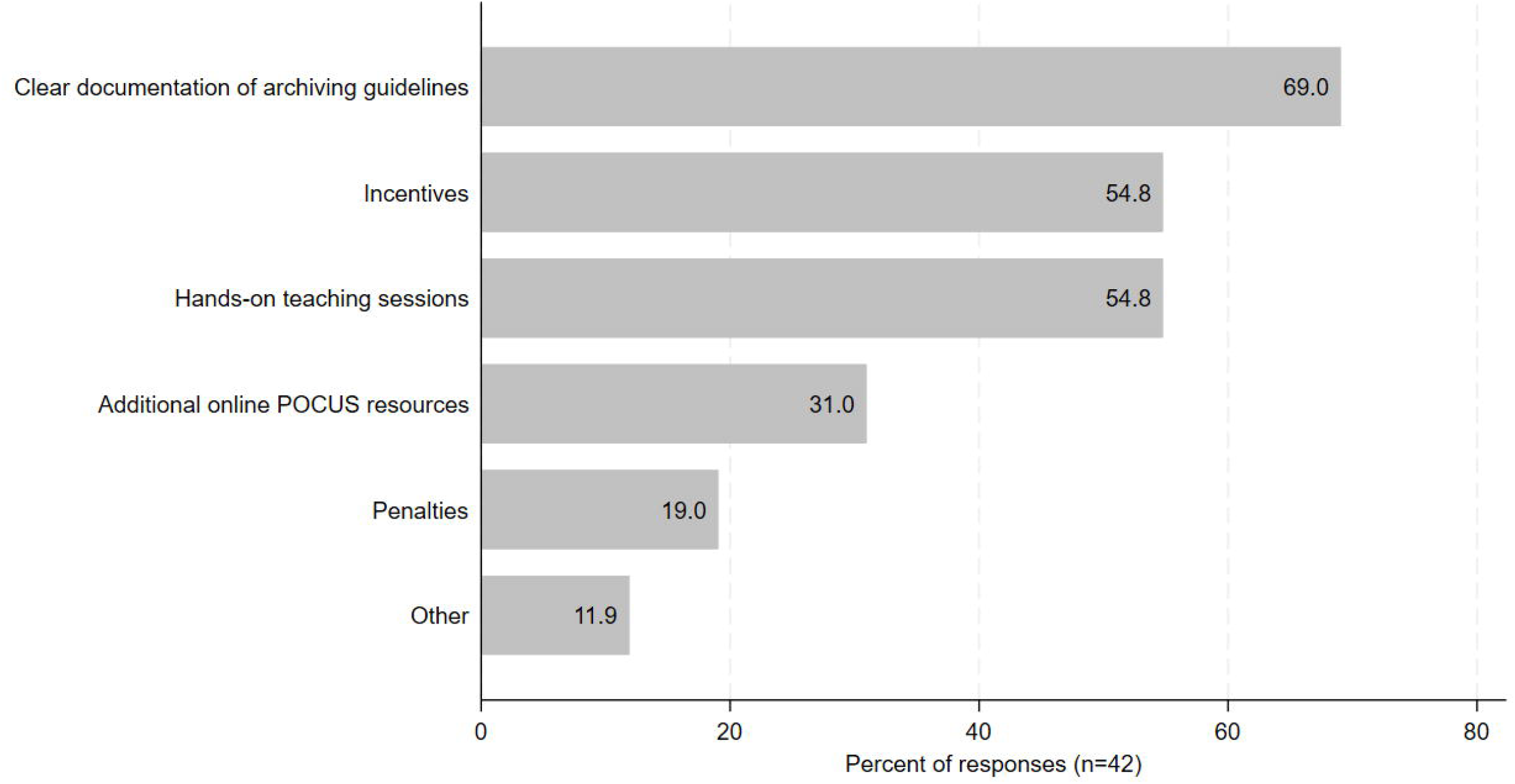
POCUS Facilitators

Perceived POCUS facilitators were *clear documentation protocols, hands-on teaching sessions, and incentives for completing scans* (all had >20 respondents). A few participants thought that additional online training and penalties for not completing a certain number of POCUS scans would also be facilitators (both had <10 respondents).(Table 2a, Figure 2b)

Most respondents felt that *POCUS* was a useful *medical tool* (41 or 95.4% agree/strongly agree) and that *it is important for residents and EM attendings to learn POCUS* (42 or 100% agree/strongly agree). As a large academic center, 23 respondents (54.8%) endorsed POCUS use often >1 time per week and 16 or 38.1% using it 1-2 times per month. Participants saved and documented only a mean of 51.34 of their clinical POCUS scans (S.D. 34.1), and they saved, documented, and completed a signed worksheet so that information would be sent to the electronic health record with a mean of 45.95 (S.D. 34.5).(Table 2b)

**Table 2b:** Participant questionnaire responses on clinical ED POCUS workflow.

| **Survey question** | **Participant response (response rate, percent)** | **Mean (S.D.); Median (IQR)** |
| --- | --- | --- |
| POCUS is a useful clinical tool | Strongly disagree (0, 0.0%)<br>Disagree (0, 0.0%)<br>Neutral (2, 4.7%)<br>**Agree (10, 23.3%)**<br>**Strongly agree (31, 72.1%)** |  |
| It is important for residents to learn POCUS | Strongly disagree (0, 0.0%)<br>Disagree (0, 0.0%)<br>Neutral (0, 0.0%) |  |
|  | **Agree (7, 16.3%)**<br>**Strongly agree (36, 83.7%)** |  |
| It is important for APPs to learn POCUS | Strongly disagree (0, 0.0%)<br>Disagree (0, 0.0%)<br>Neutral (6, 14.0%)<br>Agree (19, 44.2%)<br>Strongly agree (18, 41.9%) |  |
| It is important for attending faculty to learn POCUS | Strongly disagree (0, 0.0%)<br>Disagree (0, 0.0%)<br>Neutral (0, 0.0%)<br>Agree (6, 14.0%)<br>Strongly agree (37, 86.0%) |  |
| Perceived change in clinical POCUS use pre-/post-intervention | Significantly increased (4, 14.3%)<br>Somewhat increased (11, 39.3%)<br>Stayed the same (13, 46.4%)<br>Somewhat decreased (0, 0.0%)<br>Significantly decreased (0, 0.0%) |  |
| Self-reported number of POCUS scans done in the past 1 month | 0-5 scans (16, 37.2%)<br>6-10 scans (12, 27.9%)<br>11-15 scans (6, 14.0%)<br>>15 scans (9, 20.9%) |  |
| Percentage of total ED POCUS scans saved and documented on ultrasound machine (pre-intervention) | 0-25% (11, 26.1%)<br>26-50% (9, 21.4%)<br>51-75% (10, 23.8%)<br>>75% (10, 23.8%)<br>[Missing 2] | 51.34 (34.1); 50 (51) |
| Percentage of total ED POCUS scans saved, documented, and completed a worksheet in Butterfly cloud for EHR (pre-intervention) | 0-25% (15, 35.7%)<br>26-50% (7, 16.7%)<br>51-75% (9, 21.4%)<br>>75% (9, 21.4%)<br>[Missing 2] | 45.95 (34.51); 50 (55) |
| Percentage of total ED POCUS scans saved, documented, and completed a worksheet in Butterfly cloud for EHR (post-intervention) | 0-25% (0, 0.0%)<br>26-50% (4, 14.3%)<br>51-75% (9, 32.1%)<br>>75% (15, 53.6%)<br>[Missing 0] | [cannot calculate due to grouped responses] |
| POCUS intervention was useful | **Useful (20, 74.1%)**<br>Not useful (7, 25.9%)<br>[Missing 1] |  |
| POCUS intervention was effective | **Very useful (10, 35.7%)**<br>**Somewhat effective (6, 21.4%)**<br>Neutral (8, 28.6%)<br>Ineffective (0, 0%)<br>[Missing 0] |  |
| ED Clinical POCUS workflow knowledge assessment scores (post-intervention) | **92.9% Pass**<br>7.1% Fail<br>N=28 participants | 10 Mean score correct (0.89 S.D.)<br>10 Median score correct (1 IQR) |

\*POCUS intervention: “ultrasound faculty live demos on shift, PowerPoint/online presentation, and QR codes to reference material reviewing US machine use and Butterfly documentation”

Post-intervention, 20/28 respondents found the POCUS education/training intervention to be useful (74.1%), which included “ultrasound faculty live demos on shift, PowerPoint/online presentation, and QR codes to reference material reviewing US machine use and Butterfly documentation”. Ten (35.7%) found the training intervention very useful, 6 (21.4%) somewhat effective, 8 (28.6%) felt neutral about the intervention, and no respondents thought it was ineffective.(Table 2b)

Most participants felt comfortable or very comfortable with diagnostic POCUS both pre-intervention (76.8% performing, 72.1% teaching) and post-intervention (89.3% performing, 82.1% teaching). Even more participants felt comfortable or very comfortable with procedural POCUS both pre-intervention (88.4% performing, 83.8% teaching) and post-intervention (92.8% performing and 92.9% teaching). There were too few responses from APPs to make significant conclusions representative of their subgroup.(Table 3) Even with the higher initial reported rates, participant self-perceived comfort in performing diagnostic and procedural POCUS had a statistically significant increase from pre-to post-intervention (p=0.100, p=0.784 respectively). Self-perceived comfort in teaching also increased for procedural but not diagnostic POCUS (p=0.022, p=0.166, respectively). Mean knowledge scores on post-questionnaires were 10 (0.89 S.D.) or a score of 92.9%, with 26/28 participants passing the assessment (score >90%).(Tables 3–4)

**Table 3:** Summary comparison of clinical ED POCUS data pre- and post-intervention.

| **Self-perceived POCUS abilities (questionnaires)** | **Pre-intervention**<br>**July – December 2023** | **Post-intervention**<br>**January – June 2024** | **p-value** |
| --- | --- | --- | --- |
| Comfort performing diagnostic POCUS | Very uncomfortable (1, 2.3%)<br>Uncomfortable (2, 4.7%)<br>Neutral (7, 16.3%)<br>Comfortable (22, 51.2%)<br>Very comfortable (11, 25.6%) | Very uncomfortable (0, 0.0%)<br>Uncomfortable (0, 0.0%)<br>Neutral (3, 10.7%)<br>Comfortable (17, 60.7%)<br>Very comfortable (8, 28.6%) | 0.100 |
| Comfort teaching diagnostic POCUS | Very uncomfortable (2, 4.7%)<br>Uncomfortable (3, 7.0%)<br>Neutral (7, 16.3%)<br>Comfortable (24, 55.8%)<br>Very comfortable (7, 16.3%) | Very uncomfortable (0, 0.0%)<br>Uncomfortable (0, 0.0%)<br>Neutral (5, 17.9%)<br>Comfortable (16, 57.1%)<br>Very comfortable (7, 25.0%) | 0.022 |
| Comfort performing procedural POCUS | Very uncomfortable (1, 2.3%)<br>Uncomfortable (3, 7.0%)<br>Neutral (1, 2.3%)<br>Comfortable (15, 34.9%)<br>Very comfortable (23, 53.5%) | Very uncomfortable (0, 0.0%)<br>Uncomfortable (0, 0.0%)<br>Neutral (2, 7.1%)<br>Comfortable (13, 46.4%)<br>Very comfortable (13, 46.4%) | 0.784 |
| Comfort teaching procedural POCUS | Very uncomfortable (2, 4.7%)<br>Uncomfortable (2, 4.7%)<br>Neutral (3, 7.0%)<br>Comfortable (22, 51.2%)<br>Very comfortable (14, 32.6%) | Very uncomfortable (0, 0.0%)<br>Uncomfortable (0, 0.0%)<br>Neutral (2, 7.1%)<br>Comfortable (14, 50.0%)<br>Very comfortable (12, 42.9%) | 0.166 |
|  | **Pre-intervention**<br>**July – December 2023** | **Post-intervention**<br>**January – June 2024** | **p-value** |
| **Total Clinical ED POCUS completed:** | **1511** | **2130** | **0.0035** |
| Draft status | 671 (44.4%) | 919 (43.1%) | 0.0133 |
| Pending attending attestation | 58 (3.8%) | 175 (8.2%) | 0.0040 |
| Finalized study | 782 (51.8%) | 1036 (48.6%) | 0.0101 |
| QA reviewed study | 224 (14.8%) | 401 (18.8%) | 0.0228 |
| **Total Clinical ED POCUS by exam type:** |  |  |  |
| Aorta | 21 | 33 | 0.0988 |
| Biliary | 48 | 65 | 0.1636 |
| Cardiac | 418 | 528 | 0.0217 |
| EFAST (trauma) | 140 | 209 | 0.0148 |
| DVT (deep veins) | 8 | 18 | 0.0265 |
| Lung | 70 | 112 | 0.0154 |
| Obstetrics/gynecology | 99 | 175 | 0.0016 |
| Ocular | 22 | 25 | 0.3997 |
| Renal | 53 | 68 | 0.0985 |
| Soft tissue/<br>musculoskeletal | 54 | 98 | 0.0274 |

| **Clinical ED POCUS billing data\*\*** | **Pre-intervention**<br>**July – December 2023** | **Post-intervention**<br>**January – June 2024** | **p-value** |
| --- | --- | --- | --- |
| Total number of POCUS billed exams | 680 | 985 |  |
| Total POCUS amount billed | Approximately \$200-250,000 | Approximately \$250-300,000 | |
| Total POCUS amount reimbursed | Approximately \$25-50,000 | Approximately \$25-50,000 | |
| Monthly number of POCUS billed exams | 255 (July 2023)<br>278 (August 2023)<br>266 (September 2023)<br>232 (October 2023)<br>254 (November 2023)<br>260 (December 2023) | 295 (January 2024)<br>311 (February 2024)<br>277 (March 2024)<br>308 (April 2024)<br>351 (May 2024)<br>308 (June 2024) | 0.0008 |
| Monthly POCUS amount billed | Approximately \$60-90,000 | Approximately \$8-15,000 | 0.0439 |
| Monthly POCUS amount reimbursed | Approximately \$70-100,000 | Approximately \$10-20,000 | 0.1225 |

\*POCUS intervention: “ultrasound faculty live demos on shift, PowerPoint/online presentation, and QR codes to reference material reviewing US machine use and Butterfly documentation”
\*\*Clinical ED POCUS billing data: Dollar amounts for revenue and reimbursement are presented as approximate values due to institutional confidentiality restrictions.

The most common POCUS scans performed both pre-/post-intervention were <u>cardiac</u> (418 pre to 528 post-intervention), then <u>EFAST</u> (140 to 209), <u>obstetrics/gynecology</u> (99 to 175), and <u>lung</u> (70 to 112). The total number of POCUS scans increased from 1511 scans pre-intervention to 2130 post-intervention (p=0.0035), with 782 studies finalized pre-intervention and 1036 post-intervention.(Table 4)

**Table 4:** Post-intervention Clinical ED POCUS workflow knowledge assessment results.

| Participant ID | Training Level | Correct | Incorrect | Percent correct | Pass/Fail |
| --- | --- | --- | --- | --- | --- |
| 1 | attending | 10 | 1 | 90.9% | Pass |
| 2 | attending | 11 | 0 | 100.0% | Pass |
| 3 | attending | 8 | 3 | 72.7% | Fail |
| 4 | attending | 7 | 4 | 63.6% | Fail |
| 5 | APP | 11 | 0 | 100.0% | Pass |
| 6 | attending | 10 | 1 | 90.9% | Pass |
| 7 | resident<br>PGY2 | 10 | 1 | 90.9% | Pass |
| 8 | resident<br>PGY2 | 11 | 0 | 100.0% | Pass |
| 9 | resident<br>PGY2 | 11 | 0 | 100.0% | Pass |
| 10 | attending | 10 | 1 | 90.9% | Pass |
| 11 | attending | 10 | 1 | 90.9% | Pass |
| 12 | APP | 10 | 1 | 90.9% | Pass |
| 13 | attending | 11 | 0 | 100.0% | Pass |
| 14 | resident<br>PGY3 | 11 | 0 | 100.0% | Pass |
| 15 | attending | 10 | 1 | 90.9% | Pass |
| 16 | resident | 10 | 1 | 90.9% | Pass |
|  | PGY1 |  |  |  |  |
| 17 | resident<br>PGY1 | 10 | 1 | 90.9% | Pass |
| 18 | attending | 10 | 1 | 90.9% | Pass |
| 19 | attending | 10 | 1 | 90.9% | Pass |
| 20 | attending | 11 | 0 | 100.0% | Pass |
| 21 | resident<br>PGY1 | 10 | 1 | 90.9% | Pass |
| 22 | resident<br>PGY2 | 11 | 0 | 100.0% | Pass |
| 23 | attending | 11 | 0 | 100.0% | Pass |
| 24 | attending | 10 | 1 | 90.9% | Pass |
| 25 | resident<br>PGY1 | 10 | 1 | 90.9% | Pass |
| 26 | attending | 10 | 1 | 90.9% | Pass |
| 27 | attending | 10 | 1 | 90.9% | Pass |
| 28 | attending | 10 | 1 | 90.9% | Pass |

**Table 5:** Clinical ED POCUS scans performed in the six months pre- and post-intervention.

|  | Pre-intervention |  |  |  |  |  | Post-intervention |  |  |  |  |  |
| --- | --- | --- | --- | --- | --- | --- | --- | --- | --- | --- | --- | --- |
|  | Jul<br>-23 | Aug<br>-23 | Sep<br>-23 | Oct<br>-23 | Nov<br>-23 | Dec<br>-23 | Jan<br>-24 | Feb<br>-24 | Mar<br>-24 | Apr<br>-24 | May<br>-24 | Jun<br>-24 |
| **<u>Total POCUS completed:</u>** | **212** | **193** | **203** | **269** | **331** | **303** | **337** | **345** | **313** | **321** | **372** | **442** |
| **Draft status** | 99 | 73 | 91 | 136 | 138 | 134 | 132 | 166 | 120 | 136 | 185 | 180 |
| **Pending attending attestation** | 7 | 13 | 6 | 6 | 9 | 17 | 20 | 30 | 26 | 19 | 24 | 56 |
| **Finalized study** | 106 | 107 | 106 | 127 | 184 | 152 | 185 | 149 | 167 | 166 | 163 | 206 |
| **QA reviewed study** | 10 | 19 | 32 | 42 | 67 | 54 | 58 | 44 | 66 | 61 | 60 | 112 |
| **<u>POCUS by exam type:</u>** |  |  |  |  |  |  |  |  |  |  |  |  |
| **Aorta** | 5 | 3 | 1 | 4 | 2 | 6 | 3 | 2 | 8 | 5 | 5 | 10 |
| **Biliary** | 5 | 4 | 11 | 5 | 12 | 11 | 14 | 11 | 9 | 4 | 7 | 20 |
| **Cardiac** | 58 | 53 | 55 | 74 | 94 | 84 | 92 | 94 | 83 | 83 | 75 | 101 |
| **EFAST (trauma)** | 17 | 21 | 20 | 24 | 34 | 24 | 25 | 26 | 39 | 31 | 38 | 50 |
| **DVT (deep veins)** | 2 | 0 | 2 | 3 | 0 | 1 | 3 | 4 | 1 | 2 | 5 | 3 |
| **Lung** | 7 | 12 | 12 | 18 | 14 | 7 | 26 | 24 | 16 | 14 | 13 | 19 |
| **Obstetrics/gynecology** | 20 | 13 | 10 | 19 | 22 | 15 | 31 | 17 | 28 | 31 | 31 | 37 |
| **Ocular** | 3 | 7 | 4 | 1 | 2 | 5 | 9 | 1 | 2 | 2 | 1 | 10 |
| **Renal** | 9 | 8 | 8 | 6 | 14 | 8 | 13 | 13 | 11 | 6 | 9 | 16 |
| **Soft tissue/musculoskeletal** | 4 | 9 | 7 | 4 | 16 | 14 | 14 | 8 | 13 | 22 | 15 | 26 |
| **Total POCUS completed:** | 212 | 193 | 203 | 269 | 331 | 303 | 337 | 345 | 313 | 321 | 372 | 442 |
| **Draft status (percent)** | 46.7% | 37.8% | 44.8% | 50.6% | 41.7% | 44.2% | 39.2% | 48.1% | 38.3% | 42.4% | 49.7% | 40.7% |
| **Pending attestation (percent)** | 3.3% | 6.7% | 3.0% | 2.2% | 2.7% | 5.6% | 5.9% | 8.7% | 8.3% | 5.9% | 6.5% | 12.7% |
| **Finalized (percent)** | 50.0% | 55.4% | 52.2% | 47.2% | 55.6% | 50.2% | 54.9% | 43.2% | 53.4% | 51.7% | 43.8% | 46.6% |
| **QA reviewed (percent)** | 4.7% | 9.8% | 15.8% | 15.6% | 20.2% | 17.8% | 17.2% | 12.8% | 21.1% | 19.0% | 16.1% | 25.3% |

Many studies were still in draft status, signifying a high area of potential revenue loss both pre-intervention (671/1511 studies or 44.4%) and post-intervention (919/2130 or 43.1%). There were fewer studies pending attending attestation pre-intervention (58/1511 or 3.8%) and post-intervention (175/2130 or 8.2%) but still categorized as lost revenue. Approximately 14.8% of studies were QA reviewed pre-intervention and 18.8% post-intervention.(Table 3) The number of clinically billed scans increased from 680 to 985 pre-/post-intervention (p=0.0008). Monthly POCUS revenue also increased by approximately $55,000 in total billed (p=0.0439) and $10,000 in total reimbursed (p=0.1225).(Tables 3&5)

## Discussion

Identifying clinical POCUS use barriers and facilitators is important to implement program changes that are feasible, acceptable, and sustainable. Our study found that <u>POCUS use barriers</u> were mixed, with the most significant barriers including *time constraints on shift, forgetting to complete the worksheet on the Butterfly archiving system*, and *internet connectivity problems with losing saved images*. <u>POCUS facilitators</u> included *clear documentation protocols, hands-on teaching sessions*, and *incentives for completing scans* (all with >20 respondents agreed or strongly agreed).

As a large academic medical and trauma center, ED physicians and APPs at our site felt more comfortable performing and teaching POCUS scans, citing a lack of POCUS knowledge in performing or interpreting exams as an insignificant barrier. It is not surprising that faculty employed at an academic medical center, who are credentialed in ultrasound through residency training, do not perceive their POCUS knowledge as a barrier to use. In contrast, POCUS knowledge is still identified as a barrier at other community hospitals/EDs employing non-EM trained physicians or APPs, who typically lack POCUS training and are not as facile with its clinical use.^6,9,11,12,14^

Interestingly, many of the POCUS barriers identified in both EM attendings and residents are similar to those identified in a mixed-methods study we performed at a local VA hospital ED. At the VA, a separate EM attending group who had mixed training backgrounds (emergency, internal, and family medicine training).(Theophanous) Creation of a standardized ED POCUS documentation and archiving system at the VA site, which is reported as a POCUS facilitator in our study, increased clinical POCUS scans four-fold from 72 to 267 scans in 6 months.^14,15^ Some of these same POCUS barriers are also identified in the national VA healthcare system, as reported by Boyd and Remskar et al, with the biggest barriers being a lack of POCUS training, system infrastructure, and equipment.^11,13^ A national VA POCUS survey identified that only 61/115 VA sites are using POCUS, despite an increase in POCUS technology and utilization nationwide over the past 30 years.^10^ Furthermore, many rural or community EDs still lack POCUS equipment and training, thus expansion of this protocol to additional sites to identify and address barriers and facilitators is needed.^6–8,11^

In our study, EM attendings and residents believed POCUS documentation was already standardized and straightforward. In contrast, Schnittke’s study in EM residents noted that documenting POCUS for medical decision-making was a significant barrier to performing POCUS clinically. Thus, they incorporated resident in-person education on the POCUS documentation and archiving system.^16^ They similarly identified a need for addressing time constraints, streamlining documentation, and strengthening faculty support for resident-performed POCUS.^16^

Regarding POCUS facilitators, hands-on teaching remains the mainstay as an effective training method for POCUS education.^1,19^ It employs the medical education framework of Ericsson’s Deliberate Practice, which is learning by doing and repeating a task until it becomes familiar and second nature.^18,24–25^ Learners cycle through a process of planning, concentration/dedication, repetition/revision, and self-reflection, building knowledge from an existing foundation.^24,25^ With hands-on teaching, the learners actively participate by scanning standardized or clinical patients, under direct supervision from an ultrasound-trained faculty.^19^ Hayward et al built an EM POCUS curriculum in a Canadian hospital using Ericsson’s Deliberate Practice educational framework, increasing resident POCUS competency from 33% to 100% of PGY2 residents qualifying for the Canadian Emergency Ultrasound Society (CEUS) practitioner examination.(Hayward) The Deliberate Practice model yields excellent results but is very time consuming for ultrasound faculty and difficult to feasibly achieve at sites that may lack additional POCUS training opportunities.^18,24^

Studies and online podcasts have addressed the problem of time constraints on shift, with examples including finding the ultrasound machine at the beginning of the shift and maintaining it nearby, plugged in and ready to be used, plus bringing the machine into the patient’s room during initial assessment. Our institution has marked US machine locations within each section of the ED and a machine tracking system to save time with finding equipment in a large, busy ED. Also, residents can perform the US scans independently then show the images to attendings afterwards to expedite care while maintaining patient safety standards. Incentives for performing POCUS were favored as a facilitator, while some respondents recommended penalties for clinical POCUS non-performance as a quality metric standard. Other studies have proposed reward vs penalty systems for POCUS, as well as a tiered credentialing system for hospitalists and EM physicians.^26–28^

Creation of a standardized clinical POCUS system for EDs or hospital-wide is costly yet has potential to generate great revenue for physicians and the healthcare system.^28,29^ Other studies that have performed financial analysis of POCUS systems show significant cost savings and program effectiveness through increased POCUS scans and decreased radiology studies, as well as improved ED lengths-of-stay and hospital throughput.^28,29^

Finally, middleware software such as Butterfly cloud for POCUS archiving is an essential part of ED and hospital workflow. Although this can create problems such as connectivity issues, added time to complete POCUS worksheets, and required knowledge to operate another system, the utilization of middleware has great benefits. These benefits include a storage domain for educational scans, a safety mechanism to prevent incomplete or poor images from entering the patient’s permanent medical chart, and the ability to perform quality review and faculty credentialing. None of these tasks can be done with EHR or radiology software in a single, secure location. Thus, creation of a simple, standardized process to save time and properly document POCUS scans and interpretations is important for a successful POCUS program, as noted in the literature and by our survey results.^17,19,20,28–29^

### Limitations

The single academic center study setting limits study generalizability, but this study design was chosen to focus on making improvements specific to current barriers in our site’s ED. The convenience sample method may introduce bias but was used for quality improvement and provider inclusivity. Small sample size limits scalability and power, thus additional studies with multiple sites and larger sample sizes are needed. Extrinsic factors such as problems with ED and hospital boarding and variability in staffing models and ED personnel per day/month can affect study data results and ED workflow, which may contribute to decisions on whether to perform POCUS. For our survey data, the survey response rate was low, which may have skewed results. For example, if people were not motivated to complete the surveys, then they also may not have been motivated to perform US or participate in improving the clinical ED POCUS workflow. The questionnaires were de-identified to prevent unintended work influence and bias per IRB protocol, thus ED providers did not receive direct feedback to improve their POCUS use unless they participated during clinical shift in-person education.

### Future directions

Future studies could incorporate qualitative data collection through semi-structured interviews and focus groups to further improve POCUS processes. We measured outcomes at 6 months post-intervention for feasibility, but longer timelines of 12+ months could measure sustainability. We also are planning a targeted approach for ED physicians with the highest clinical hours and lowest POCUS scans, conducting in-person individualized learning sessions to further improve clinical ED POCUS use and documentation. Finally, protocol employment at multiple different sites can elucidate common or differing POCUS use barriers and facilitators and tailor specific quality improvement initiatives based on findings.

## Conclusions

We identified barriers and facilitators to clinical POCUS use at a single site academic ED and incorporated findings into a clear standardized documentation and archival protocol. Using a multi-faceted approach with in-person teaching, an accessible reference document, and image-based learning tool, we achieved a statistically significant increase in clinical ED POCUS use and revenue post-intervention. Future studies on individualized interventions for low POCUS users or institutional-wide initiatives with clinical champions are needed to determine effects on improved patient care.

## Data Availability

All data produced in the present study are available upon reasonable request to the authors.

## Statements and declarations

The manuscript has not previously been published and is not under consideration in the same or substantially similar form in any other journal.

## Data availability

“The data that support the findings of this study are available from the corresponding author upon request and are included in the supplemental materials. The review protocol is included in the methods.”

This study was determined exempt by the Institutional Review Board (Pro00114618).

## Supplementary Files

S1: POCUS questionnaire participant comments on barriers and facilitators

S2: POCUS post-intervention questionnaire and knowledge assessment

S3: STROBE guidelines for prospective study

S4: Online PowerPoint slides from clinical POCUS training intervention

S5: Standardized POCUS reference document

