## Supplementary material for "Prospective Cohort Study Identifies Barriers to Point-of-care Ultrasound use in an Academic Emergency Department": Supp File 1

Supplemental File: Descriptive comments and Bar Graphs from Clinical ED POCUS workflow surveys

**a) Participant comments on POCUS barriers:**

**PGY1:** “Not enough ultrasound machines throughout the department. It can often take some time to locate an available ultrasound machine in the department. The current machines that we have lose battery extremely quickly and are often not ready to use, or may lose battery at the end of the scan, causing the images to be lost. Other barriers include confirmatory imaging being ordered in triage, so there is not much benefit to additional scanning even though if I were to want to answer pointed questions before a pt got additional scans, I would want to turn to my ultrasound first.”

**Attending physician:** “Encouraging residents to clinically integrate POCUS into their workflow as opposed to thinking of it as a "separate procedure". Also encourage residents to complete worksheets on time.”

**PGY2:** “no endovaginal probe”

**Attending physician:** “I find that it is not culturally common that POCUS should be done for all SOB and CP patients. Some residents have not had their Butterfly access, which has also been a barrier. Lastly, I often have difficulty finding a machine when I need one.”

**Attending physician:** “The entire process needs to be simplified to the level that it takes almost no extra time to save and interpret images after obtaining them”

**APP:** “residents/APP US must be supervised or, at least, images reviewed with attending; would otherwise do less phantom. To bill I need faculty to review; adds an extra 2 minutes to move US from speed to pod and THEN waiting for faculty to be available for review. This process could be more streamlined if butterfly upload was faster, allowing for remote review (provider in speed, attending in pod).”

**Attending physician:** “culture of residents telling attending when they are doing US, I will be there. but after the fact, I can't teach as well or effectively”

**Attending physician:** “Time required to fill out US worksheets in addition to other documentation, necessity to access/log into separate software; time/steps required to use the ultrasound tracking software”

**Attending physician:** “more hands on practice for faculty”

**Attending physician:** “Availability of ultrasounds”

**Attending physician:** “None that I can think of”

POCUS Barriers per participant pre-questionnaire responses (n=42 respondents)

|  |  |
| --- | --- |
| time constraints while on shift | 30 |
| not knowing how to save images on the ultrasound machine | 11 |
| forgetting to fill out the worksheet on Butterfly cloud after the scan is complete | 24 |
| Images/study are not uploaded to Butterfly by the end of the shift | 24 |
| not understanding the current documentation protocol | 16 |
| residents and/or APPs not completing study worksheets | 15 |
| faculty not signing completed studies | 14 |
| inability to locate the ultrasound machine | 20 |
| do not know how to use the machine tracker system (RTLS Activate or Cetani) | 18 |
| nursing or other staff are using the diagnostic machines for US-IV placement (e.g. GE, Sonosite M-turbo, or Sonosite Edge) | 6 |
| residents/APPs are performing 'phantom' scans and not saving their images | 19 |
| problems with the ultrasound machine malfunctioning | 21 |
| internet connectivity problems and losing saved images | 26 |
| difficulties with using Butterfly cloud software (not familiar with the software or not being able to find scans) | 11 |
| losing access to sign non-associated Butterfly scans after the 2-week time window | 12 |
| hospital faculty credentialing does not include all necessary specific diagnostic and therapeutic exam types (e.g. EFAST, aorta, renal, central lines) | 4 |
| ready availability of radiology-performed ultrasounds (24/7 US technician/radiologist presence at academic medical center) | 16 |
| lack of confidence in obtaining POCUS images | 9 |
| lack of confidence in interpreting POCUS images | 11 |

**b) Participant comments on POCUS facilitators (*Other facilitators in bar graph):**

**PGY3:** “Machines need to be more available and functional”

**PGY2:** “I really don't like having to complete the butterfly worksheets. I feel like there's a better way or additional ways to document”

**Attending physician:** “Improved IT/tech performance to ensure timely image upload”

**Attending physician:** “better availability of machines, upload time faster”

**PGY2:** “Ultrasound machine that doesn't lose connection”

POCUS Facilitators per participant pre-questionnaire responses (n=42 respondents)

|  |  |
| --- | --- |
| Incentives (competition, monetary bonus for faculty, more stringent faculty credentialing process) | 23 |
| Penalties for not completing a certain number of signed POCUS scans per month | 8 |
| Hands-on teaching sessions | 23 |
| Additional online POCUS resources | 13 |
| Clear documentation of archiving guidelines | 29 |
| Other (please specify) | 5 |
