## Supplementary material for "Prospective Cohort Study Identifies Barriers to Point-of-care Ultrasound use in an Academic Emergency Department": Supp File 2

**ED Provider POCUS Post-intervention Questionnaire**

June 2024

Please complete the survey. Your responses will be kept confidential. Group de-identified data will be evaluated and disseminated for ED point-of-care ultrasound (POCUS) clinical process improvements.

Name: _________________

NetID: _____

Age: _____

Gender: (Male, Female, Nonbinary, or prefer not to answer)

Ethnicity: (Hispanic/Latino/x/Spanish origin or Non-Hispanic/Latino/x/Spanish origin, or prefer not to answer)

Race: (American Indian or Alaska Native, Asian, Black or African American, Native Hawaiian or Other Pacific Islander, White, other, or prefer not to answer)

What is your level of training? (PGY1, 2, 3, fellow, attending physician, APP)

Have you completed a fellowship? If so, which? Yes	No	_____________________

Number of years in clinical practice working in the emergency department (including residency/fellowship)? _________

--How many point-of-care ultrasound exams have you personally performed in medical/PA school and residency?

a) 0-50

b) 51-100

c) 101-150

d) >150

--How comfortable do you feel **performing** POCUS for:

a) Diagnostic scans **after** the POCUS training intervention?

*Very uncomfortable, uncomfortable, neutral, comfortable, very comfortable*

b) Ultrasound-guided procedures **after** the POCUS training intervention?

*Very uncomfortable, uncomfortable, neutral, comfortable, very comfortable*

--How comfortable do you feel **teaching** others how to perform:

a) A diagnostic POCUS scan while working in the ED?

*Very uncomfortable, uncomfortable, neutral, comfortable, very comfortable*

b) Ultrasound-guided procedures in the ED?

*Very uncomfortable, uncomfortable, neutral, comfortable, very comfortable*

Did you find the ultrasound educational/training intervention useful? (i.e. ultrasound faculty live demos on shift, PP/online presentation, and QR code on the US machines and in the pods to reference material reviewing US machine use and Butterfly documentation)

a) Yes
b) No

Did you find the ultrasound intervention teaching format to be effective?

a) Very effective

b) Somewhat effective

c) Neutral

d) Somewhat ineffective

e) Very ineffective

Which component of the ultrasound intervention was useful to you? _______________

Since the POCUS intervention (6 months ago), how often do you use point-of-care ultrasound in the clinical setting?

a) Often (>1 time per week)

b) Somewhat often (1-2 times per month)

c) Occasional (<1 time per month)

d) Rarely (1-2 months per year)

e) Never

Since the POCUS intervention (6 months ago), do you feel like your ultrasound use:

a) Increased
b) Stayed the same
c) Decreased

--What percentage of your total POCUS scans have you saved AND completed the Butterfly worksheet?

a) 0-25%
b) 25-50%
c) 50-75%
d) >75%

--What are the most common types of POCUS scans that you have done in the past one month?

______________________________________________________________________

What additional type of training do you prefer to be proficient in acquiring and interpreting point-of-care ultrasound images? Please select **ALL** that apply.

A. Hands-on sessions
B. Classroom didactics
C. Web-based teaching/online modules
D. Review of ultrasound images with expert (QA sessions)
E. None (the initial training intervention was sufficient)

What else could be improved for our current clinical POCUS system? ___________________________

Please provide any additional feedback or suggestions for future training interventions. ________________________________________________________________________________

**Brief knowledge assessment:**

1) How can you more easily and most efficiently locate a missing ultrasound machine in the Duke ED?
2) What are two acceptable ways to associate or link a patient to your POCUS study?
3) What button do you need to press on the ultrasound machine to make sure that your images will be saved to Butterfly and so that the machine is ready for the next patient?
4) How do you know if your US machine is connected to the internet network?
5) What is required to perform and document a **clinical** ED POCUS study?
6) What is required to perform and document an **educational** ED POCUS study (for resident ultrasound requirements)?
7) On Butterfly cloud, how do you assign yourself to a study?
8) On Butterfly cloud, how do you associate a patient with a study?
9) On Butterfly cloud, what step allows your study to be sent to Epic MaestroCare with a generated ultrasound report?
10) On Butterfly cloud, which folder must be opened after log in to find your clinical POCUS study?
11) How many days do you have to complete and sign your study in Butterfly cloud before the patient disappears from the clinical worklist?

Answers:

A. Walk around the entire department searching for the appropriate ultrasound machine
B. **Log into the RTLS Activate (FindIT) tracker system to find the US machine location in the ED**
C. Take the US machine from the resuscitation bays for your study in the Pods
D. Search for the US machine location in the ED using Butterfly Cloud
A. Find the patient in the “worklist” on the US machine and click on their name prior to starting your exam
B. Type in the MRN and the study will be automatically associated
C. Type in the patient’s name or MRN on the machine and then find the patient on the Butterfly cloud on the computer and click “Associate”
D. **A & C**
3) A. Machine power off button

B. Patient worklist button

C. Save image button

**D.** **End exam button**

4) **A. An icon will appear in the corner of the screen (3 bars with antenna)**

B. The battery icon will have an electric bolt over it

C. The power button will light up green

D. An icon will appear in the corner of the screen (two boxes with an arrow connecting them)

5) A. A resident performs an exam on a patient who has a pending CT scan order for comparison and documents the study on their own

B. **A resident performs an exam on their own then shows the images to the clinical attending afterward and completes the Butterfly worksheet**

C. An APP performs an exam and documents the study in Butterfly on their own

D. A resident performs an exam while the clinical attending is in the room and documents it in Epic without completing the Butterfly worksheet

6) A. **A resident performs an exam on a patient who has a pending CT scan order for comparison and documents the study on their own**

B. A resident performs an exam on their own then shows the images to the clinical attending afterward and completes the Butterfly worksheet

C. An APP performs an exam on a patient who has a pending chest x-ray and documents the study in Butterfly

D. A resident performs an exam while the clinical attending is in the room and completes the butterfly worksheet with attending signature

7) **A. Type your net user ID (initials and numbers) or your name into the “authors” box on the left side of the screen**

B. Type your patient’s MRN into the “authors” box on the left side of the screen

C. Type your password into the “authors” box on the right side of the screen

D. Once the patient’s name and MRN are associated with the study, you do not need to add your name to sign/finalize the study

8) A. Click on the patient exam to open it, then type your net user ID (initials and numbers) into the “choose from worklist” box on the left side of the screen

**B. Click on the patient exam to open it, then at the top center of the screen click on the study and click “choose from worklist” to find the patient by name or MRN**

C. Type your patient’s MRN into the “authors” box on the left side of the screen

D. Once your name is added to the study, you do not need to associate the patient’s name and MRN to sign/finalize the clinical study

9) A. The exam is finalized and sent to the clinical chart in Epic when the user completes the exam worksheet

B. The exam is finalized and sent to the clinical chart in Epic when the resident clicks on “sign final”

**C. The exam is finalized and sent to the clinical chart in Epic when the attending physician clicks on “sign final”**

D. The exam is finalized and sent to the clinical chart in Epic when the user clicks “end exam” on the US machine with an active internet connection

10) A. Using the search bar at the top of the screen, search for “clinical archive”

B. In butterfly cloud on the lefthand side of the screen, click on “educational archive”

**C. In butterfly cloud on the lefthand side of the screen, click on “clinical archive”**

D. Using the search bar at the top of the screen, search for “educational archive”

11) A. 24 hours

B. 48 hours

C. 1 week or 7 days

**D. 2 weeks or 14 days**
