## Supplementary material for "Prospective Cohort Study Identifies Barriers to Point-of-care Ultrasound use in an Academic Emergency Department": Supp File 4

**Interventions:**

- Intervention 1: PowerPoint format photos with descriptions to facilitate use of each of the cart-based ultrasound machines in the Duke ED (Venue, M-turbo). The goal is to have a readily available PowerPoint to serve as a reminder for how to use the worklist to attach a patient to your study, tag yourself to the study to help locate the study in butterfly, and how to use butterfly to fill out the corresponding worksheet for your study. This will also include a few pointers for how to troubleshoot the machines.

The power point will include:

- Images of the worklist (confirm the correct day for overnight shifts)
- Images of the Wi-Fi icon (connected to Hearts)
- Image of where to type your net ID
- Image of the spooler to show the status of your study (sent/pending/failed)
- Images of the butterfly interface to help locate your drafts, find studies you are not tagged to in the clinical archive, and review how to fill out a sample worksheet
- Intervention 2: Small group (1-4 people) in-person review during clinical shift or pre/post-shift change of the ultrasound machines to demonstrate how to use the worklist, tag yourself to the study and go through troubleshooting the machines. Additionally, practice one study (clinical or educational), and go through the corresponding butterfly worksheet.
  - Review the GE Venue/M-turbo worklist/query and have each person practice using the worklist to attach a patient and themselves as the reading doctor
  - Practice one scan together (either clinical or educational) to review ‘knobology’ and labeling as needed
  - Practice troubleshooting Wi-Fi on the machine and demonstrate how a hard reset would be done if the machine is not functioning properly
  - Practice locating and completing one worksheet in butterfly
  - Practice locating a scan in butterfly ‘clinical archive’ that you did not assign yourself to
- Intervention 3: A short online handout review of the most common ED clinical POCUS uses and the associated common acceptable indications and required views accessed via QR codes placed on the ultrasound machines and ED pod whiteboards

**Approach to interventions:**

- Plan A: Distribute Intervention 1 (PowerPoint) to all faculty and residents after the Pre-Survey is completed, and simultaneously send a link for sign-up dates/times to complete Intervention 2. Any of the ultrasound faculty/fellow may participate in Intervention 2, using a standardized approach to reviewing the machines and completing worksheets in Butterfly. Targeting pre- and post-shift sign-out times may be an effective time to maximize participation. Coordinating ahead of time with residents/attending may help increase total participation. The goal timeframe for Intervention 2 is 4-8 weeks from the date Intervention 1 is distributed.
- Plan B: Discuss time at faculty meeting and resident conference to review Intervention 1. Use a portion of SIM/ultrasound conference time to complete Intervention 2 with the residents. Teach faculty during quarterly faculty development sessions to complete Intervention 2 with the EM attendings and APPs. Use a sign-up link with dates/times for attendings or request an in-person individualized review with one of the ultrasound faculty (for pre- or post-shift sign-out and during clinical shifts).
