## Supplementary material for "Prospective Cohort Study Identifies Barriers to Point-of-care Ultrasound use in an Academic Emergency Department": Supp File 5

**Ultrasound Required Views for Logging/Credentialing**

-**Incidentalomas** **Reminder**: if you find incidental abnormals (eg renal cysts with septations that are suspicious, aorta top end of normal but not abnormal – talk with your attending to decide how to inform patient/whether they should have follow-up sono or other imaging)

-**Educational Scans:** You should always inform a patient when performing an exam for educational purposes, say something such as “Hello, I am an ED resident/attending performing ultrasound for educational purposes. I will share my findings with your treating doctors. Would this be ok with you?”. It may also help to mention this will not be billed or added to their chart.

**1. Aorta –**

- Abdominal aorta from diaphragm to bifurcation of iliacs. Should taper distally (normal diameter is <3cm outer to outer wall)

- **Required Views for Credit:**

1. Still images x2 (1 suprarenal, 1 infrarenal) of transverse axis aorta with	calipers measuring diameter

2. Video clips of aorta in longitudinal and transverse axis

3. Optional/additional: mid aorta view

- Look for: diameter >3cm, dissection flap, intraluminal thrombus, etc.

**- Points for your interpretation**

- note suprarenal and infrarenal measurements and whether there is appropriate tapering

**-** state if aneurysm present or if dissection flap present

**2. Cardiac**

-**Required Views for Credit:** **Clip each of…**

1. Parasternal long (dot to R shoulder, visualize RV, LA, LV, MV, AV, Aortic root)

2. Parasternal short (dot to R hip, visualize LV, RV, and papillary muscles)

3. Apical 4 Chamber (at apex, visualize RA, RV, LA, LV, TV, MV)

4. Subxiphoid (visualize RA, RV, LA, LV)

5. Optional/additional: IVC measurement during sniff/respirat

-Look for: pericardial effusion/tamponade, signs of decreased systolic function, signs of R heart strain, IVC assessment for fluid tolerance

**-Points for your interpretation**

- presence of pericardial effusion? If so, tamponade?

- estimate of EF (if reduced, mild/moderate/severe?; is there focal hypokinesis?)

- IVC diameter, collapsibility (> or <50%) both clip and M mode

**3. FAST or EFAST**

-assessing for presence of pericardial effusion, free intraperitoneal fluid, +/- hemo/pneumothorax

-**Required Views for Credit: Clip each of…**

1. Subxiphoid

2. RUQ (don’t forget to visualize caudal tip of liver, fluid hides here!)

3. LUQ (try to get look above spleen and below diaphragm, fluid hides here!)

4. Suprapubic (ideal to look in both transverse and sagittal planes)

5. Lungs (if doing EFAST) – may also include M-mode view of lung line

-Look for: pericardial effusion, free fluid in abdomen, pleural effusion/hemothorax, absence of lung sliding, lung point, or “bar code sign” to suggest pneumothorax

**-Points for your interpretation**

**-** Is free fluid present in abdomen? In which view(s)?

- Is pneumothorax and/or hemothorax present? In which lung(s)?

- Is pericardial effusion present? If so, is tamponade present?

**4. Biliary**

-Normals:

--Gallbladder Wall Thickness (measure anteriorly!) <0.3cm

--Common bile duct (measure inner to inner wall): 3mm at age 30 + 1mm per decade of life thereafter. (*Tip on finding CBD:* find portal vein (PV) in long view, with probe in transverse orientation. On top of PV is CBD, also in long view. Can measure here, or to get short view: Rotate probe 90° to see Mickey Mouse sign 🡪 PV = face; hepatic artery = medial ear; CBD = lateral ear. CBD should have NO flow on color.)

-**Required Views for Credit**

1. Clip of GB in transverse axis

2. Clip of GB in long axis

3. Clip of GB with patient in LLD position (if stones present)

4. Measurement of anterior GB wall thickness (best measured adjacent to liver)

5. If possible – CBD diameter measurement

-Look for: gallstones. Presence of GB wall thickening/pericholecystic fluid/sonographic Murphy’s sign 🡪 all signs of cholecystitis. CBD dilation and/or stones in CBD.

**-Points for your interpretation**

- presence of gallstones?

-signs of cholecystitis? Comment on each

-CBD diameter

**5. Renal**

-**Required Views for Credit**

1. Clip of each kidney in long axis, can add transverse to better visualize renal pelvis

2. Clips of bladder in transverse and longitudinal

-Look for: Hydronephrosis (mild/moderate/severe), cysts, bladder thickening, stones or clots, etc.

**-Points for your interpretation**

- presence of hydronephrosis? If yes, to what degree?

-any obvious stones or other findings like cysts, clot

**6. Transabdominal OB**

-Full bladder makes best sonographic window

-**Required Views for Credit: Clip of each…**

1. Uterus in long axis

2. Uterus in transverse axis

3. R adnexa in transverse

4. L adnexa in transverse

5. Fetal cardiac activity: clip of or rate measured in M-mode (no Doppler!!!)

6. Gestational age measurement (CRL if <12 weeks, BPD if >12 weeks)

-Look for: An intrauterine pregnancy, as defined by a gestational sac WITH AT LEAST a yolk sac inside. Then look for fetal pole +/- cardiac activity depending on age (usually 5-6 wks on transvaginal, 6-7 wks transabdominal). Presence of free fluid in pelvis. Ovarian cysts (>5cm more likely to torse but cannot *rule out* torsion by size)

**-Points for your interpretation**

- Is IUP present? If so, dates and FHR if available

- If no IUP, is there free fluid, adnexal masses? Also consider proceeding to transvaginal study for better visualization

-Gestational age: if BPD limited, can obtain FL/AC/HC

**Additional imaging (not considered core indications)**

**7. Transvaginal OB**

-You should always do a transabdominal scan first. Then have patient empty bladder for TVUS.

-**Required Views for Credit: Clip of each of the following…**

1. Uterus in Sagittal/Long axis

2. Uterus in Coronal/transverse axis

3. R adnexa in transverse

4. L adnexa in transverse

5. If fetal pole seen, measure for gestational age (CRL <12wks, BPD >12wks)

6. If fetal pole seen, try to take measure FHR in M-mode (NO Doppler), or at least take clip of fetal cardiac activity

-Look for: An intrauterine pregnancy, as defined by a gestational sac WITH AT LEAST a yolk sac inside. Then look for fetal pole +/- cardiac activity depending on age (usually 5-6 wks on transvaginal). Presence of free fluid in pelvic. Ovarian cysts (>5cm more likely to torse, but cannot *rule out* torsion by size)

**-Points for your interpretation**

- Is IUP present? If so, dates and FHT if available

- If no IUP, is there free fluid, adnexal masses?

**8. DVT**

-**Required Views for Credit**

1. Clip each with compression of: CFV, SFV, popliteal vein

2. Clip with augmentation at CFV when squeeze calf with color Doppler on

-Look for: noncompressible vein (must fully collapse), clot in lumen, sluggish or incomplete flow with augmentation) – do NOT compress if you see obvious thrombus!

**-Points for your interpretation**

- is DVT present? If so at what level?

**9. Thoracic/Lung**

-**Required Views for Credit**

1. Clip at apex of each lung to evaluate for lung sliding (can include M-mode)

2. Clip at inferolateral base of each lung to evaluate for effusion

3. Clips with adequate depth (≥10cm) to evaluate for B-lines

4. Optional/additional: Clips of any area to evaluate for consolidation/pneumonia

-Look for: absence of lung sliding, lung point, or “bar code sign” for pneumothorax; fluid collection/effusion; focal B-lines, hepatization, air bronchograms, and/or shred sign for pneumonia; >3 B-lines per intercostal space in multiple lung regions bilaterally for pulmonary edema (can also see this with ILD/pulmonary fibrosis)

**-Points for your interpretation**

- are there signs of pneumothorax? Which lung(s)?

- are there B-lines? If so, are they focal or diffuse?

- is there a pleural effusion? Which lung(s)?

- are signs of pneumonia seen on US? If so, where?

**10. Abscess**

-**Required Views for Credit**

1. Clips through the region of interest in two planes

2. Clip showing effects of compression (may see pus swirling)

3. Clip with color to look for vascularity as with mass, lymph node, or pseudoaneurysm-Look for: presence of fluid collection, cobblestoning (interstitial edema), “swirl sign” of purulent material, absence of internal vascularity, size/depth of collection if present

**-Points for your interpretation**

- is an abscess/fluid collection present?

- is there associated blood flow to suggest pseudoaneurysm or other vascular structure?

**11. Musculoskeletal**

Watch out for anisotropy – if probe is not perpendicular to tendon/ligament, may lose signal and appear anechoic because all sound waves are reflected away

-**Required Views for Credit**

1. Clips through the region of interest in two planes

2. Optional/additional: Clips of pathology (tendon rupture, fracture, etc.)

Look for: disruption of cortex (fracture) or discontinuity of tendon, fluid collection in the joint or around a tendon (tendonitis/tenosynovitis), dislocation

**-Points for your interpretation**

- is there a fracture or tendon/ligament injury?

- is there a dislocation? If reduction attempted, is the joint reduced?

- is there a joint effusion? (can guide arthrocentesis)

**12. Ocular**

Be sure to increase the gain sufficiently to view the posterior chamber

-**Required Views for Credit**

1. Clips through each eye in both sagittal and axial planes

2. Clips with probe static and eye looking back and forth

3. Optional/Additional: Measurement of optic nerve sheath diameter (must measure across ONS 3mm deep to retina for accurate measurement)

Look for: vitreous hemorrhage (“washing machine” sign), posterior vitreous detachment, retinal detachment, increased ONSD (<5mm normal, 5-6mm indeterminate, >6mm abnormal)

**-Points for your interpretation**

- is there vitreous hemorrhage? PVD? RD? If RD, is the macula off?

- is the ONSD increased?

**All residents are required to have 180 total scans by the end of residency. This is broken up into 25 scans each of the six core topics (Aorta, Cardiac, FAST/EFAST, Biliary, Renal, TA-OB) plus 30 other scans of any type (e.g. thoracic, soft tissue/MSK, DVT, procedural, etc.)**
